# A retrospective investigation of the population structure and geospatial distribution of *Salmonella* Paratyphi A in Kathmandu, Nepal

**DOI:** 10.1101/2024.01.08.23300021

**Authors:** Elli Mylona, Pham Thanh Duy, Jacqueline Keane, Sabina Dongol, Buddha Basnyat, Christiane Dolecek, Phat Voong Vinh, Nga Tran Vu Thieu, To Nguyen Thi Nguyen, Abhilasha Karkey, Stephen Baker

## Abstract

*Salmonella* Paratyphi A, one of the major etiologic agents of enteric fever, has been on the rise over the last decades in certain endemic regions compared to *S.* Typhi, the most prevalent cause of enteric fever. Despite this, data on the prevalence and molecular epidemiology of *S.* Paratyphi A remain scarce. Here, we analysed the whole genome sequences of a total of 216 *S*. Paratyphi A isolates originating in Nepal between 2005 and 2014, of which 200 were from acute patients and 16 from chronic carriers of enteric fever. By using the recently developed genotyping framework for *S*. Paratyphi A (paratype), we identified several genotypes circulating in Kathmandu. Importantly, we observed an unusual clonal expansion of genotype 2.4.3 over a four-year period that spread geographically and replaced other genotypes. This rapid genotype replacement is hypothesised to have been driven by both reduced sensitivity to fluoroquinolones and genetic changes to virulence factors, such as functional and structural genes of type 3 secretion systems. Finally, we show that person to person is likely the most common mode of transmission and chronic carriers play a limited role in maintaining disease circulation.

## Introduction

Enteric fever is a life-threatening systemic infection caused by *Salmonella enterica* serovars Typhi (*S.* Typhi) and Paratyphi A (*S.* Paratyphi A), B and C. Enteric fever poses a major public health burden in countries with poor sanitation, poor food handling and limited access to clean water. There are an estimated 11–27 million cases annually and >120,000 associated deaths globally [1]. Incidence is higher among children, teenagers, and young adults, and transmission occurs via the faecal-oral route with contaminated food or water [1,2]. Symptoms typically include fever, headache, anorexia, and/or constipation or diarrhoea [2]. In addition, a fraction of enteric fever cases (1–5%) in endemic regions can progress into chronic carriage, which is typically characterized by colonisation of the gallbladder, clinical clearance but shedding of bacteria in faeces, and is thought to be an ecological niche promoting bacterial adaptation and disease transmission [3,4].

While *S*. Typhi is responsible for most enteric fever cases, *S.* Paratyphi A isolation rate has been on the rise in the recent years causing about a third of global cases, and >40% of cases in certain countries, such as India and Nepal [1,5,6]. However, sparse epidemiological data, inappropriate diagnostic tools and poor surveillance in several endemic countries underestimate the actual incidence of *S*. Paratyphi A and there is limited understanding of the population structure, spatiotemporal spread, and transmission dynamics. To this end and aligning with efforts towards recording and standardizing the genotyping of *S*. Typhi, a single nucleotide polymorphism (SNP)-based tool was recently developed to assign genotypes to *S*. Paratyphi A organisms called ‘Paratype’ [7]. While the symptomatic disease caused by *S*. Typhi and *S*. Paratyphi A is clinically indistinguishable [8], the two organisms are thought to have differential epidemiology, with *S*. Paratyphi A infection more likely to occur outside the household as opposed to *S*. Typhi [9]. The development of tools such as paratype [10], promise to improve understanding of *S*. Paratyphi A epidemiology and genomic surveillance.

Notably, and unlike *S*. Typhi, there are currently no licensed vaccines targeting *S.* Paratyphi A, but several are in development [11]. The most likely antigen for a vaccine is the *S.* Paratyphi A O-antigen (O2); however, some isolates have been found to harbour SNPs in the O-antigen encoding region, which now also form part of the Paratype output in order to keep track of such changes [10]. It remains unclear whether nonsynonymous SNPs in this region may lead to changes in the O2-antigen structure that could potentially complicate vaccine-induced antibody binding.

Unlike *S*. Typhi, reports of multi-drug resistance (MDR; resistance to ampicillin, contrimoxazole, and chloramphenicol) in *S*. Paratyphi A are rare [12–14]. However, the emergence of resistance to key antimicrobials used for treatment is increasingly being reported, and most *S*. Paratyphi A organisms have reduced susceptibility to nalidixic acid and ciprofloxacin, due to mutations in the quinolone resistance-determining region [15–19]. Additionally, sporadic cases of ceftriaxone resistance, as well as azithromycin resistance have also been reported in the Indian subcontinent [20–22]. Antimicrobial resistance (AMR) becomes a larger problem in areas with poor sanitation and food hygiene, namely areas where *S.* Paratyphi A is endemic and, therefore, incidence reporting and AMR detection becomes crucial for managing enteric fever.

South Asia is a hotspot for *S.* Paratyphi A infections, with Nepal being a highly endemic area of enteric fever [1]. While there has been an overall decline in the incidence of enteric fever in the last decades, the prevalence of *S.* Paratyphi A isolation in Nepal rose from 23 to 34% of *Salmonella* cases between 1993 and 2003 [14], and remained high between 2005 and 2009 at 31.5% of total enteric fever cases [23]. Furthermore, 1.6% of patients undergoing cholecystectomy were found to be carrying *S.* Paratyphi A organisms in their gallbladder [24]. There is a limited understanding of the population structure of *S.* Paratyphi A and the genetic relatedness between isolates from acute cases and carriers. Here, we retrospectively studied 216 *S.* Paratyphi A organisms, both acute and carrier, isolated in an urban hospital in Kathmandu, Nepal, between 2005 and 2014 by whole genome sequencing (WGS). We determined the structure, genetic diversity, and dynamics of this population, both in a local and a global context. We observed a clonal expansion of one genotype, and thus further investigated the spatial spread of the isolates and identified changes in virulence factors that may explain this expansion. Our study offers new insights into the population structure and dynamics of *S*. Paratyphi A in this highly endemic region.

## Materials and methods

### Ethics

Samples were obtained from studies that were approved by the institutional ethical review boards of Patan Hospital, The Nepal Health Research Council, and The Oxford University Tropical Research Ethics Committee. All enrolees were required to provide written informed consent for the collection and storage of all samples and subsequent data analysis. In the case of those under 18 years of age, a parent or guardian was asked to provide written informed consent.

### Study site

Bacterial isolates were obtained from blood cultures obtained from patients enrolled on clinical trials (see below) conducted at Patan hospital in Lalitpur Sub-Metropolitan City (LSMC), which is one of the three urban districts that constitute the Kathmandu Valley. The Bagmati River separates the LSMC from the Kathmandu Metropolitan City at the North and West sides of the city. According to the 2001 Nepalese census, it has an area of 15.43 km^2^ and is divided into 22 municipal wards with a population of 126,991 people living in 68,922 households with an average household size of 4.66 persons. At the time of sampling most people living in the area were poor and used municipal water and traditional stone spouts as the main water sources. Patan Hospital is the only hospital in LSMC with 320 beds and provides treatment for about 320,000 outpatients and 20,000 inpatients every year.

### Bacterial isolates

A total of 223 *S*. Paratyphi A isolates were collected from a wide range of studies conducted at Patan hospital, Nepal between 2005 and 2014; from these, 206 blood isolates were collected from four randomized control trials: gatifloxacin vs cefixime (2005) [25], gatifloxacin versus chloramphenicol (2006-2008) [26], gatifloxacin versus ofloxacin (2008-2011) [27], gatifloxacin versus ceftriaxone (2011-2014)[28][28] and a matched case-control study (2011) [23]; 17 chronic isolates was recovered from a chronic gallbladder carriage study (2007-2010) [24].

All isolates originated from acute enteric fever patients were recovered from blood cultures collected at the time of enrolment. Briefly, blood samples (5–10 mL for adults; 1–3 mL for children) were inoculated into media containing tryptone soya broth and sodium polyanethol sulphonate to a total volume of 50 mL. The collecting vials were incubated at 37 °C and examined daily for bacterial growth for 7 days. Positive samples were sub-cultured on MacConkey agar and suspected *Salmonella* colonies were identified using standard biochemical tests and serotype-specific antisera (Murex Biotech, Dartford, England).

Chronic isolates were collected from stool and bile extracts from patients undergoing cholecystectomy. Bile samples were inoculated in Selenite broth and Peptone broth overnight before being sub-cultured onto MacConkey and Xylene Lysine Deoxycholate agar. *Salmonella* colonies were identified using API20E (bioMerieux, Inc) and confirmed using the same serotype-specific antisera as above.

Epidemiological and clinical information was collected from the above studies where available. Major reported variables included age, sex, main water source(s), water treatment method(s), clinical symptom(s) at presentation, antimicrobial resistance information and individual GPS location.

The minimum inhibitory concentrations (MICs) of nalidixic acid, ofloxacin, ciprofloxacin, gatifloxacin, azithromycin, chloramphenicol, amoxicillin, cotrimoxazole and ceftriaxone were determined by E-test (AB Biodisk, Solna, Sweden) according to the manufacturer’s instructions. MICs were interpreted by using the 2020 CLSI guidelines [29].

### Genome sequencing

Genomic DNA was extracted using the Wizard Genomic DNA Extraction Kit (Promega, Wisconsin, USA) and 2 μg of DNA was subjected to indexed-tagged paired-end sequencing on an Illumina HiSeq 2000 platform to generate 100 bp paired-end reads following the manufacturer’s recommendations.

Seven genomes were excluded from analyses due to low quality scores based on abnormal genome size (<4 or >5 million base pairs), >1000 contigs, positive status for contamination using confindr [30], and/or low score in bactinspector (available at: https://gitlab.com/antunderwood/bactinspector), resulting in a final total number of 216 isolates, of which 200 acute isolates and 16 carrier isolates (Supplementary Table S1). Similar quality screening criteria were applied to the global collection (see below).

### Phylogenetic analysis

Sequencing reads were mapped to *S*. Paratyphi A AKU12601 (accession: FM200053) reference genome using the nf-core bactmap pipeline v1.0.0 (https://nf-co.re/bactmap/1.0.0; [31]) to make pseudogenomes based on high quality positions in VCF files. In all analyses, *S*. Typhi CT18 (accession: GCF_000195995.1_ASM19599v1) was used as an outgroup; the sequence was shredded using fasta2fastqshredder.py (https://github.com/sanger-pathogens/bact-gen-scripts) to mimic the data format of the sequenced isolates. Using the pseudogenome multi-alignment file obtained from bactmap, repetitive regions, prophages, and recombinant regions were removed using remove-blocks (https://github.com/sanger-pathogens/remove_blocks_from_aln) and a pre-defined coordinates list specific for Paratyphi A (PARAREPEAT; https://github.com/katholt/typhoid) followed by a screen and removal of recombinant regions with Gubbins v3.2.0 [32]. The multi-sequence alignment file obtained from gubbins was fed into RAxML-ng v1.1.0 [33] to infer a maximum-likelihood phylogenetic tree using a GTRG+gamma model and the autoMRE convergence test that resulted in 750 bootstrap pseudo-analyses of the alignment. For the global collection analysis, following file selection based on quality checks as above, 828 publicly available *S*. Paratyphi A sequences were used ([34], Supplementary Table S2) and repetitive regions, prophages and recombinant regions were excluded as above. The filtered Nepali collection and global collection alignment files were combined and used to run Gubbins. The maximum-likelihood phylogeny tree was inferred as above with 200 bootstraps. Outgroup rooting was performed in FigTree v1.4.4 (https://github.com/rambaut/figtree) and trees were visualized and annotated using iTOL [35]. SNP distance matrices were produced using snp-dists v0.8.2 (https://github.com/tseemann/snp-dists) using the Gubbins output files.

### Genotyping, antimicrobial resistance genes, plasmid sequences, and virulence factors

Using the raw sequencing reads, or assemblies for the global collection, genotypes were assigned using the Paratype genotyping tool [7], which also detects mutations in *gyrA*, *parC*, *acrB*, and O-antigen region genes (*rmlB*, *rmlD*, *rmlC*, *ddhB*, *prt*, *wzx*).

Using SRST2 v.2.0.2 [36], known AMR genes were detected with CARD [37] and ARGannot [38] databases, and plasmid replicons were detected with the PlasmidFinder [39] database. Different alleles and/or mismatches in virulence genes were screened using SRST2 and the Virulence Factor Database (VFDB) [40]. Genes that had a difference (either allele or mismatch) within more than 20% of organisms in each clade were considered for visualisation.

### Clustering analysis

Precise GPS and collection date data were used to detect spatiotemporal clusters were using a Space Time Permutation model [41] in SatScan™ v10.1.2 (available at: https://www.satscan.org) at the month level with 2005-06-17 as start study date and 2014-12-01 as end study date and using the high/low rate detection and default settings.

### Data visualization and statistical analysis

When needed, data were formatted using the tidyverse v2.0.0 [42] and dplyr v1.1.2 [43] in R [44]. Statistical analysis and graphs were produced in Graphpad Prism v9.5.1 and Rstudio v2023.06.0+421 (Posit software, PBC) using ggplot2 v3.4.2 [45] and maps were annotated using leaflet v2.1.2 (https://CRAN.R-project.org/package=leaflet) and captured using webshot v0.5.4 (https://CRAN.R-project.org/package=webshot).

## Results

### Epidemiological and clinical observations

The genomes of 216 *S.* Paratyphi A isolates originating from studies conducted in Patan Hospital in Lalitpur within the Kathmandu valley, Nepal, were whole genome sequenced. Organisms included in this analysis were isolated from patients presenting with acute enteric fever (referred to as enteric fever hereafter) (200/216) [23,25–28] or from the bile/gallbladder of patients undergoing cholecystectomy (i.e., chronic carriers and referred to as carriers hereafter; 16/216) [24]. As dictated by the design of the clinical studies contributing these organisms, most patients were resident within a 2.5 km^2^ radius from Patan Hospital, while carriers were more dispersed (Figure 1A). Most acute enteric fever patients suffered from headache (92.5%; 185/200), with anorexia being the second most common symptom reported (68.5%; 137/200) (Figure 1B). More than a third of patients reported being nauseous (37%, 75/200) and/or having abdominal pain (32.5%, 65/200); diarrhoea (13.5%, 27/200) and constipation (12%, 24/200) were less common (Figure 1B). A fraction of patients reported none of these symptoms (2.5%, 5/200). The median age of enteric fever patients was 20 (IQR 13–24), while the median age of carriers was 38 (IQR 33–43.5) (see also Figure S1). Most enteric fever patients were male (72%; 144/200), while less than a third were female (28%; 56/200). In comparison, a quarter of carriers were male (4/16) and the majority were female (68.8%, 11/16); this information was missing for one patient. Collectively, the representation of *S.* Paratyphi A from acute enteric fever patients spanned a nine-year period, from 2005 to 2014. During this time, cases peaked in the summer months (June–August; total of 126/200), while remained relatively low in the winter months (October–February; total of 17/200, Figure 1C), which is comparable to previous observations in this area [46].

**Figure 1.**
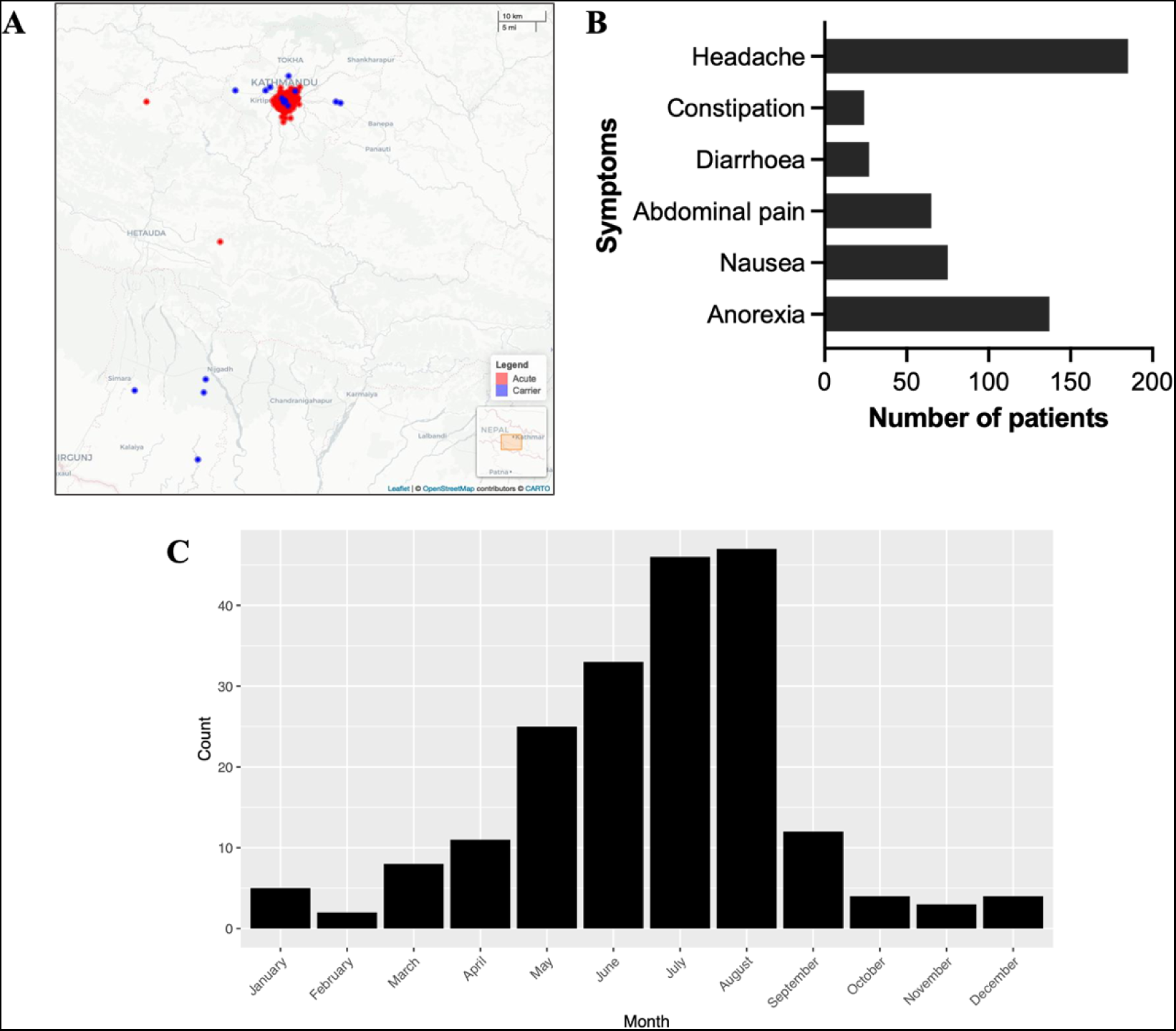
Epidemiological observations of *S*. Paratyphi A patients in Nepal. (A) Geographic map of Nepal showing the precise residence location of acute enteric fever patients (red) and carriers (blue). (B) Numbers of acute enteric fever patients (200) reporting symptoms of headache, constipation, diarrhoea, abdominal pain, nausea, and/or anorexia. (C) Monthly distribution of acute enteric fever cases during 2005–2014.

### S. Paratyphi A population structure in Nepal

To explore the structure of this *S*. Paratyphi A population, we inferred a whole genome phylogeny based on 1,082 SNPs and assigned genotypes based on the recently developed paratype scheme [7] (Figure 2 and Supplementary Table S1). As a reference, the lineages of the old genotyping scheme [5] are also shown in Supplementary Table S1. Overall, *S*. Paratyphi A isolates were highly related, having an average pairwise difference of only 10 SNPs (IQR 8–90).

**Figure 2.**
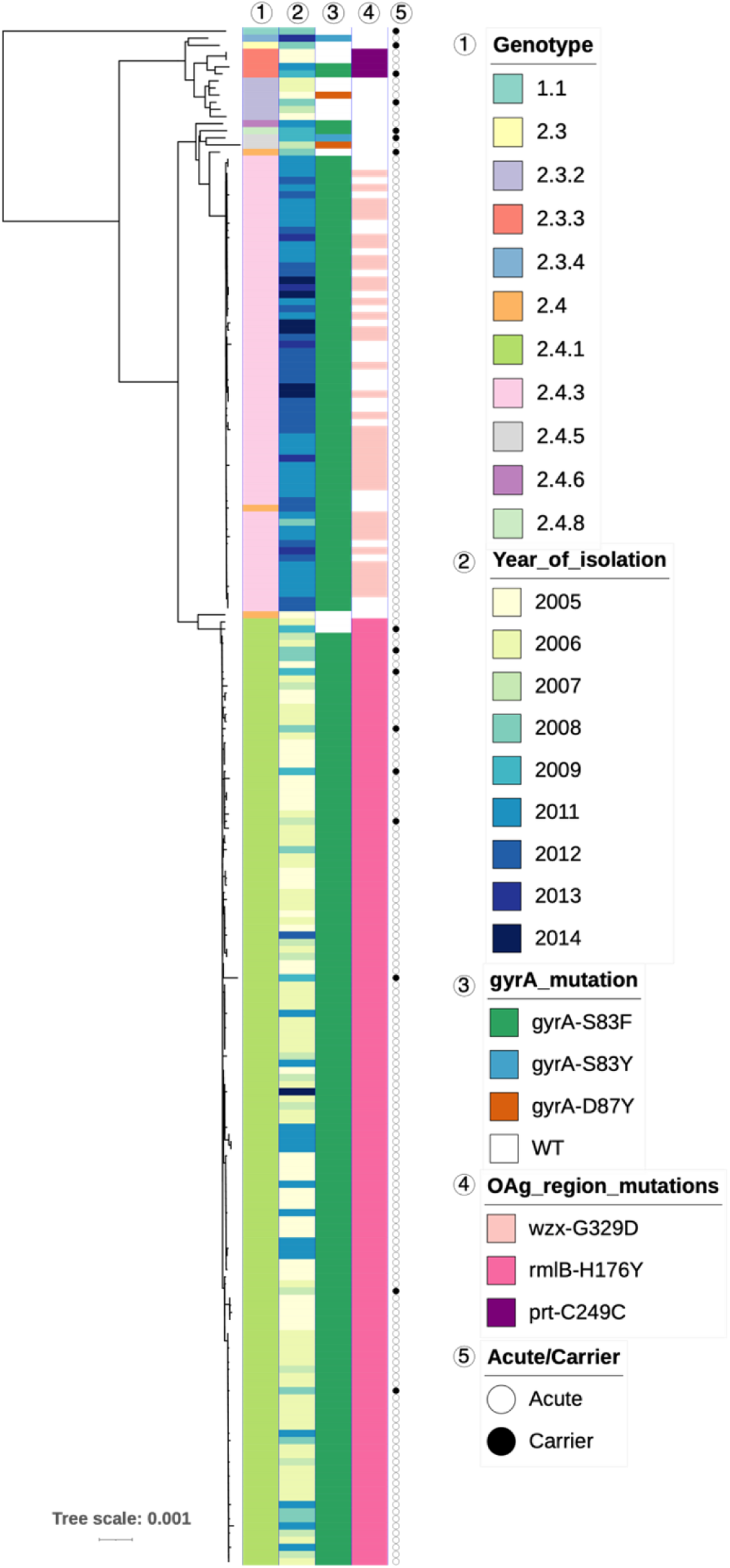
The population structure of *S.* Paratyphi A in Nepal in 2005–2014. Rooted SNP-based maximum likelihood phylogenetic tree of 200 acute and 16 carrier *S*. Paratyphi A isolates from Nepal. Annotations are marked with a number on top that corresponds to the numbers on the key. Tree was constructed with the reference strain included, which was removed at the visualisation stage.

The most common genotype in the Nepali *S.* Paratyphi A population was 2.4.1 (61.6%; 133/216), while the second most common was 2.4.3 (29.2%; 63/216). Genotypes 2.3.2 (2.8%; 6/216), 2.3.3 (1.9%; 4/216), 2.4 (1.4%; 3/216), and 2.4.5 (0.9%; 2/216) were present in small numbers, while 2.3, 2.3.4, the newly defined 2.4.6 and 2.4.8, as well as 1.1 were each recorded once (0.5%; 1/216 each).

The *S*. Paratyphi A population in Nepal split largely into two major subclades, comprised mainly of isolates belonging to genotype 2.4.3, that clustered closely to a small group of 2.4 organisms, and genotype 2.4.1 (Figure 2). In line with the genotyping scheme, genotypes belonging to the 2.3 secondary clade of Paratype (2.3, 2.3.2, 2.3.3, 2.3.4) clustered mainly in a separate, smaller clade. Although the number of carrier *S.* Paratyphi A isolates in this collection was small, these belonged mostly to 2.4.1 (9/16), as well as 2.4 (1/16), 2.4.5 (1/16), 2.4.8 (1/16), 2.3 (1/16), 2.3.2 (1/16), 2.3.3 (1/16), and 1.1 (1/16), the latter being the most distantly related to the rest. The distribution of genotypes over age in enteric fever patients is shown in Figure S1; we did not observe any correlation between the age brackets and genotype (Pearson’s chi squared test *p*>0.05). Furthermore, we visualised the isolates by genotype on a geographical map of the area (Figure S2A-C) and used precise GPS and collection date information to perform spatiotemporal analysis. A space-time permutation model predicted two statistically significant clusters; one high-rate cluster in a 1.66 km radius containing 20 isolates (*p*=0.00024) and one low-rate cluster in a 0.49 km radius containing 33 isolates (*p*=0.047) (Figure S2D). Other statistically not significant clusters (*p*>0.05) were also observed (Figure S2D). Notably, we observed a variety of genotypes within these clusters.

The two major clades observed, that of genotype 2.4.1 and that of genotype 2.4.3, were each highly clonal with isolates of genotype 2.4.3 differing by 2 SNPs on average (IQR 1–3) upon pairwise comparison; isolates of genotype 2.4.1 differed by a median of 7 SNPs (IQR 6–8). Overall, the carrier isolates carried on average 16 SNPs (IQR 14–92) when compared to acute isolates, but, as suggested by the assigned genotypes, carrier isolates were closely related to acute cases that shared the same genotype, such as that genotype 2.4.1 carrier isolates differed by 6 SNPs (IQR 5–8). The distantly related isolate of genotype 1.1 showed a pairwise difference of 438 SNPs with all other isolates (IQR 436–438) irrespective of whether they were from acute or carrier cases.

Most organisms isolated in the first half of the study period (2005–2009) belonged to genotype 2.4.1 with a few cases caused by *S*. Paratyphi A belonging to 2.3.2, 2.3.3, 2.4 or 2.4.5 genotypes, while genotype 2.3 was sampled only in 2008 (Figure 2 and 3). In the second half of the study period (2011–2014), we observed that genotype 2.3 and 2.3.2 were not represented. Additionally, genotype 2.3.3 organisms were diminished, and there was an extensive decrease in isolation of genotype 2.4.1 organisms. Conversely, there was a substantial increase in the isolation of organisms belonging to genotype 2.4.3, which accounted for most organisms isolated during this time period (Figure 3). A limited number of 2.3.4 organisms were identified in 2013 only. The temporal distribution was also supported by the pairwise SNP difference within the clades, such that 2.4.1 had accumulated more SNPs.

**Figure 3.**
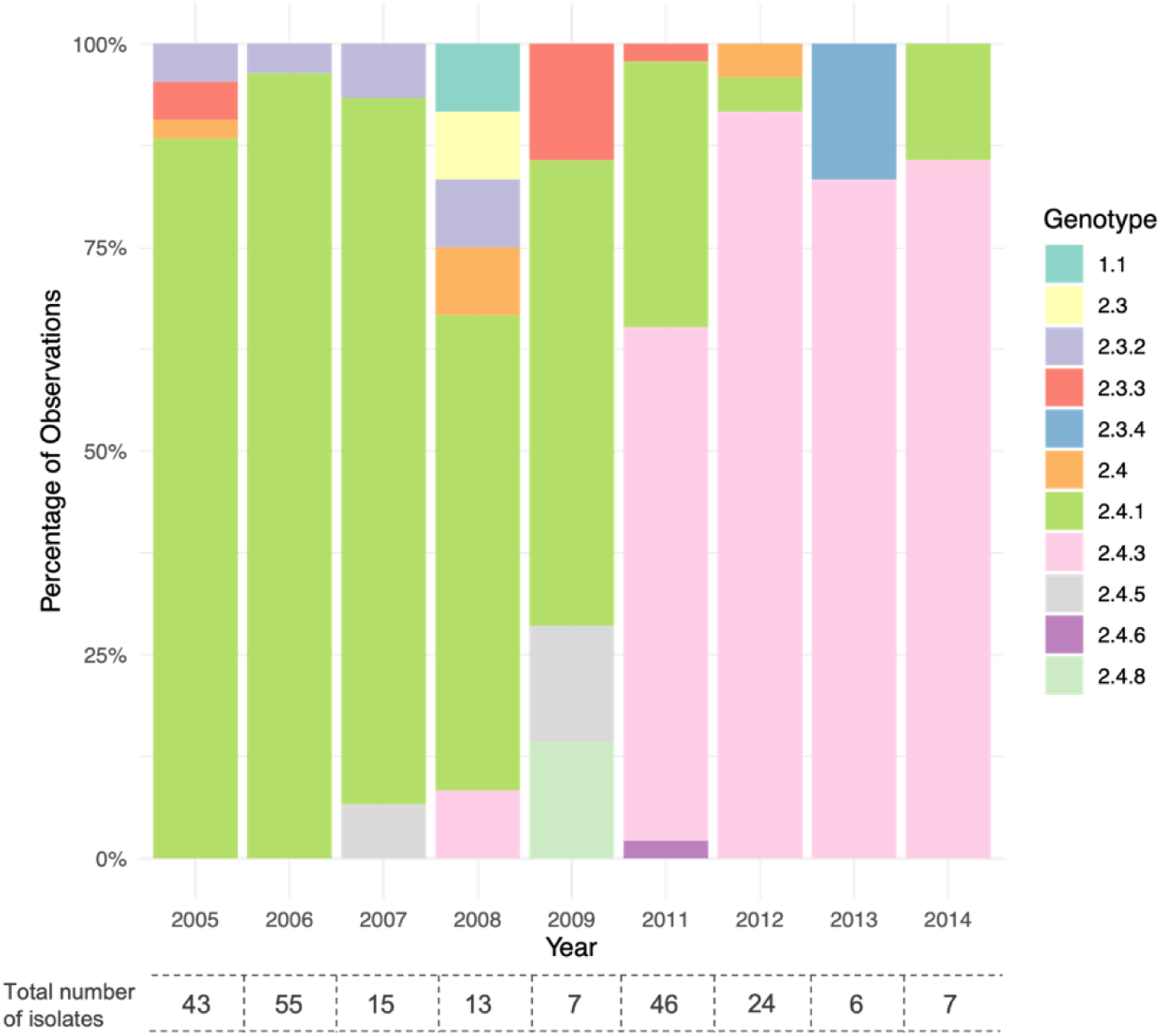
Annual distribution of *S.* Paratyphi A genotypes in Nepal, 2005-2014. Stacked bar plot showing the percentage of each genotype in each year. No isolates sampled in 2010. Numbers below bar correspond to the absolute total numbers of isolates in each year.

### Water source and water treatment methods in acute cases

Water source has been previously linked to enteric fever outbreaks and transmission, with the contamination of water supplies with sewage waste and general sanitation issues likely responsible for enteric fever in areas like Kathmandu [47,48]. We had information regarding the main water source and water treatment method for 156 acute cases. From those, more than half (82/156; 52.6%) used the municipal water supply, while smaller numbers used bottled water, water from wells, stone spout water, tanker water or a mix of these (Table 1). A third of patients did not treat their water in any way (55/156; 35.3%), another third filtered their water prior to consumption (54/156; 34.6%), while less than a fifth boiled their water (23/156; 14.7%) and very few cases chlorinated, used another way of treatment, or did not treat water at all. For each water source, filtering and no treatment were the more common treatment methods, while other methods (chlorine, boiling, other) were associated with fewer cases (Table 1).

**Table 1.** Main water source and treatment method of 156 Nepali *S.* Paratyphi A patients.

| Main water source | Count (%) | Water treatment method | Count (%) |
| --- | --- | --- | --- |
| Municipal | 82/156 (52.6) | Boiling | 12/82 (14.6) |
|  |  | Chlorine | 3/82 (3.7) |
|  |  | Filter | 38/82 (46.3) |
|  |  | Mix | 8/82 (7.8) |
|  |  | Other | 1/82 (1.2) |
|  |  | Untreated | 19/82 (23.2) |
|  |  | Unknown | 1/82 (1.2) |
| Stone spout | 23/156 (14.7) | Boiling | 4/23 (17.4) |
|  |  | Chlorine | 1/23 (4.4) |
|  |  | Filter | 3/23 (13.0) |
|  |  | Mix | 3/23 (13.0) |
|  |  | Untreated | 12/23 (52.2) |
| Bottled | 17/156 (10.9) | Boiling | 1/17 (5.9) |
|  |  | Filter | 5/17 (29.4) |
|  |  | Mix | 1/17 (5.9) |
|  |  | Other | 1/17 (5.9) |
|  |  | Untreated | 9/17 (53.0) |
| Well | 17/156 (10.9) | Boiling | 4/17 (23.5) |
|  |  | Chlorine | 1/17 (5.9) |
|  |  | Filter | 5/17 (29.4) |
|  |  | Mix | 1/17 (5.9) |
|  |  | Other | 1/17 (5.9) |
|  |  | Untreated | 5/17 (29.4) |
| Tanker | 2/156 (1.3) | Untreated | 2/2 (100) |
| Mix | 15/156 (9.6) | Boiling | 2/15 (13.3) |
|  |  | Chlorine | 1/15 (6.7) |
|  |  | Filter | 3/15 (20) |
|  |  | Other | 1/15 (6.7) |
|  |  | Untreated | 8/15 (53.3) |

Although the group of the 156 *S.* Paratyphi A cases for which we had water source and treatment information did not contain comparable numbers of genotypes, there was no correlation between water source and genotype (genotypes 2.3.2, 2.3.3, 2.3.4, 2.4, 2.4.1, 2.4.3, 2.4.5, 2.4.6; Chi squared *p*=0.248) (Figure 4). There were a comparable number of *S.* Paratyphi A of genotype 2.4.3 and genotype 2.4.1 associated with people reporting access to municipal water, stone spout and tanker water, while 2.4.3 was more common in those reporting using bottled water and 2.4.1 in those reporting using well water (Figure S3A,C). The majority of cases were associated with municipal water and hence the greater variety of genotypes identified (2.3.3, 2.4, 2.4.1, 2.4.3, and 2.4.6) (Figure S3A,C). The low numbers of *S*. Paratyphi A organisms detected following reporting chlorinated, unknown or ‘other’ treatment were all genotype 2.4.1 (Figure S3B, D), while multiple genotypes were detected even after reporting of boiling, filtering or not treating water (Figure 4, S3B,D).

**Figure 4.**
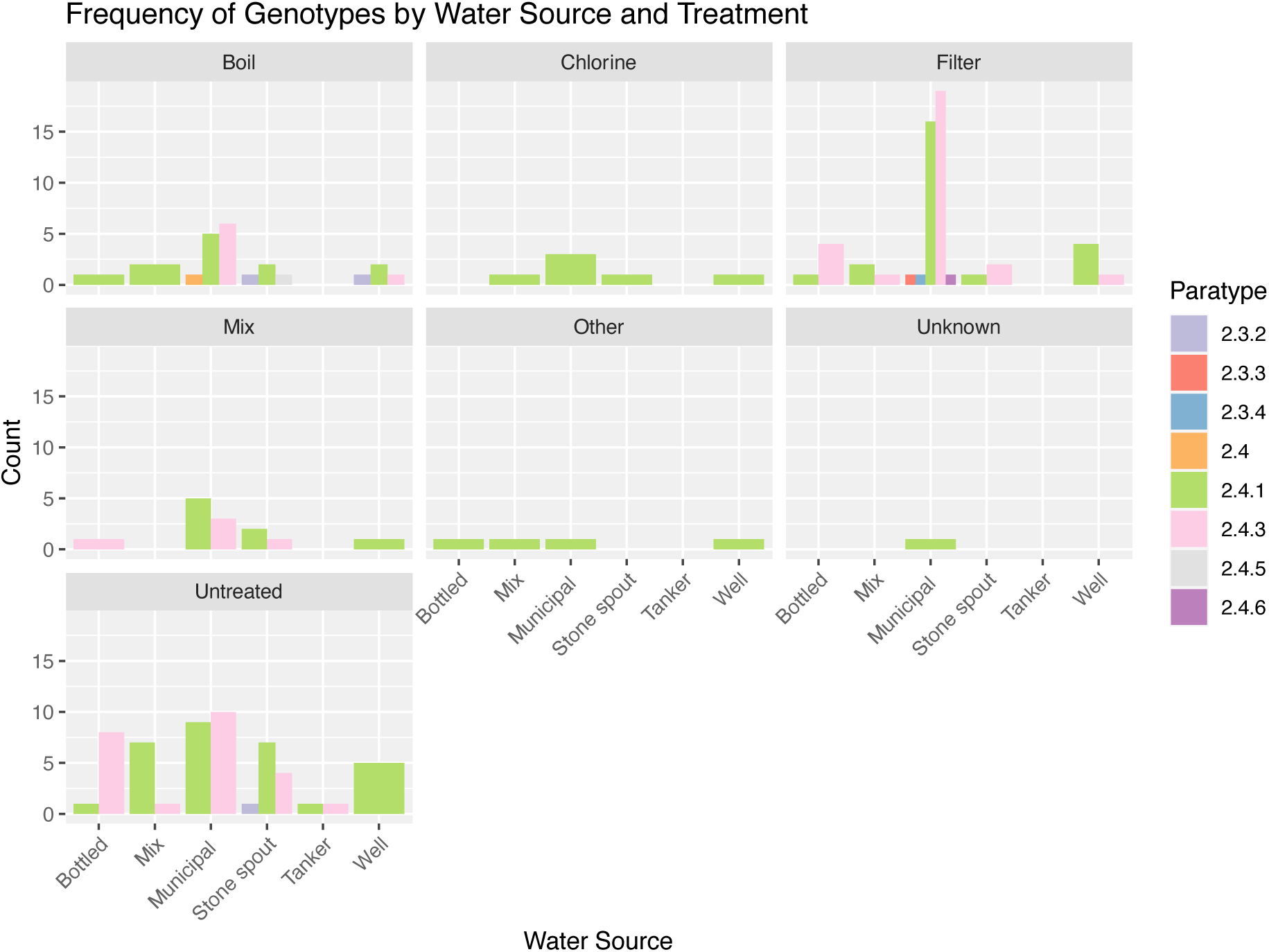
Frequency of each genotype in each water source, grouped by treatment method. Bar plots showing the absolute numbers of each genotype per water source for each treatment method.

### The Nepali S. Paratyphi A population in a global context

To investigate the *S*. Paratyphi A population dynamics in a global context, we constructed a global phylogeny based on 6,811 SNPs by including an additional 828 publicly available *S.* Paratyphi A genomes from a wide range of geographical regions and chronological periods (Figure 5). Genomes from our collection clustered not only with those of the same genotype in the global phylogenetic tree, but also with those originating from the same period (2005–2014) and more recently (2015–2019). In the context of this study, the more recent (2011–2014) 2.4.3 genotype clade clustered with other organisms of the same genotype and was closely related to a clade containing 2.4.2 and 2.4.4 organisms from more recent years; all these genomes originated from countries bordering Nepal; India, Pakistan, and Bangladesh. The majority of organisms from our collection clustered closely to organisms originating from countries within South and Southeast Asia.

**Figure 5.**
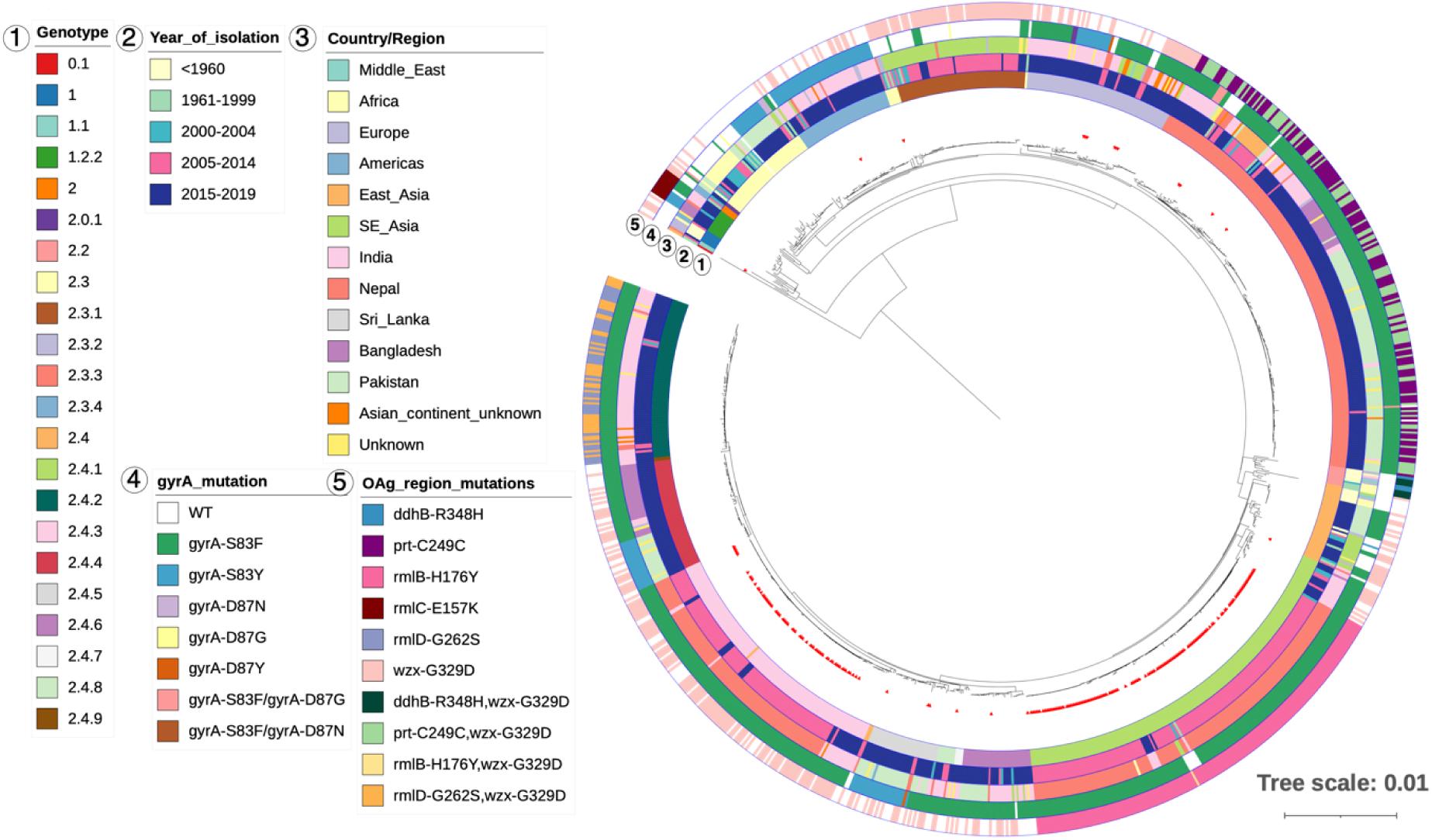
The Nepali *S.* Paratyphi A collection in a global context. Rooted SNP-based maximum likelihood phylogenetic tree of 828 global isolates and the 216 Nepali isolates (marked with red arrows). Annotation rings are marked with a number that corresponds to the numbers on the key. NB the tree was re-rooted on midpoint on the longest branch for visualisation purposes. Tree was constructed with the reference strain included, which was removed at the visualisation stage.

### Antimicrobial susceptibility

The isolates were phenotypically tested for their antimicrobial susceptibility. All acute isolates were susceptible to amoxicillin, cotrimoxazole, ceftriaxone, and gatifloxacin. The majority of acute isolates were susceptible (MIC ≤ 8 μg/ml; 80.6%; 87/108) or exhibited intermediate resistance (MIC 8–32 μg/ml; 18.5%; 20/108) against chloramphenicol and azithromycin (susceptible MIC ≤16 μg/ml 90.1%; 136/150 and intermediate resistance MIC 16–32 μg/ml 6.7%; 10/150, respectively). It is worth noting that the small number of acute isolates displaying resistance to azithromycin had MICs of 32– 48 μg/ml. Similarly, the majority of carrier isolates (15/16) were susceptible to chloramphenicol and/or azithromycin. Conversely, 94% of acute isolates tested (141/150) were resistant to nalidixic acid (MIC >256 μg/ml), while we recorded 14.7% (22/150) resistant (MIC >1 μg/ml) and 79.3% (119/150) intermediate resistance (MIC 0.5–0.12 μg/ml) to ciprofloxacin, as well as 48% (72/150) resistant (MIC >2 μg/ml) and 46.7% (70/150) intermediate resistance (MIC 0.25–1 μg/ml) to ofloxacin. Overall, 62.5% (10/16) of carrier isolates were resistant to nalidixic acid, which was significantly lower than acute isolates (Fisher’s exact test *p*=0.0009). Significantly fewer (Fisher’s exact test *p*<0.05) carrier isolates were ciprofloxacin-resistant (12.5%; 2/16) or intermediate (50%; 8/16) compared to acute isolates.

We also evaluated the presence of genetic markers associated with AMR in the Nepali collection of *S*. Paratyphi A. Mutations in *gyrA* are associated with reduced susceptibility to fluoroquinolones in *S*. Paratyphi A [18]. The majority of *S.* Paratyphi A isolates carried a mutation in *gyrA*; the most common mutation was a substitution of serine (S) to phenylalanine (F) at position 83 (S83F; 199/216; 92.1%), while only two isolates carried a serine to tyrosine (Y) substitution at the same position (S83Y; 0.9%) and two carried an aspartic acid (D) to tyrosine substitution at position 87 (D87Y; 0.9%) in *gyrA* (Figure 2). Notably, none of the isolates carried more than one mutation in *gyrA*, unlike more distantly related global isolates that were studied here (Figure 5). We did not find any mutations in *parC* that has previously been linked to fluoroquinolone resistance [18]. Resistance to azithromycin is an increasing problem in typhoid treatment, with a few azithromycin resistant cases reported in *S.* Paratyphi A [20,21,49]. However, we did not detect any mutations in *acrB* that are commonly associated with this phenotype [21]. All samples also carried the cryptic *aac6’-Iy* gene, which, however, is generally silent and is only expressed after rare transcriptional modifications [50].

### Genetic differences in virulence determinants between clades

To identify genetic markers associated with the expansion of either 2.4.1 or 2.4.3 clade, we next investigated the presence of plasmids, differences in gene content, and/or differences in virulence factors. By screening the collection against the PlasmidFinder database (see Methods), we found only three isolates carrying a plasmid, two genotype 2.4.3 organisms (alleles ColBS512_1_NC_010656, and Col156_1_NC_009781) and one genotype 2.3.4 organism (allele CP036184_1_p5025970) that we have previously found circulating in India [34]. We did not identify any other known plasmid replicons in this collection.

An important virulence factor for *S*. Paratyphi A is the O-antigen of the lipopolysaccharide (LPS). While O-antigen has important roles in bacterial physiology and virulence, there is added interest due to its use as a target antigen in several vaccines in development against *S*. Paratyphi A [11]. For this reason, the paratype genotyping tool also records SNP mutations for genes in the O-antigen encoding region and three of such mutations were detected in our collection (*wzx*-G329D, *rmlB*-H176Y, *prt*-C249C) (Figure 2). Interestingly, genotype 2.4.1 isolates commonly had a *rmlB*-H176Y mutation, while the *wzx*-G329D substitution was only found in 2.4.3 isolates; the frequency of these mutations did not change with time. It is worth noting that several different O-antigen gene mutation profiles were observed in a global context as well (Figure 5).

We lastly inspected the collection for differences in other virulence factors, as defined by the Virulence Factor database [40]. We identified several differences in genes associated with the *Salmonella* pathogenicity island (SPI) 1 and 2 to be associated with the 2.4.3 clade (Figure 6). More specifically, changes in SPI-2 genes included a SNP in the SPI-2 effector protein *sifA* that was associated with 2.4.3 clade and its closely related isolates (2.4, 2.4.5, 2.4.6, 2.4.8) and was not found in the rest of the collection. Similarly, clade 2.4.3 exhibited a higher rate of carrying distinct alleles for *iacP* and the SPI-1 effector protein genes *sopA* and *sopD2*, while two different *sopE* alleles clustered with each of the 2.4.3 and 2.4.1 clade. A similar pattern was observed for several SPI-2 structural/effector protein genes (*ssaJ*, *ssaN*, *ssaP,* and *steA*), as well as *sinH* that is involved in bacterial adhesion. Finally, we observed higher chance of differences in the 2.4.3 clade for a small number of fimbria-associated genes (*bcfC*, *csgA*), while this clade was more likely to lack a Type 6 secretion system (T6SS)-associated gene (STM0283). The genetic diversity identified in these virulence factors included either small mismatches (e.g. *sifA*) or larger scale changes (where different alleles carry multiple mismatches/indels, like in the case of *sopE*).

**Figure 6.**
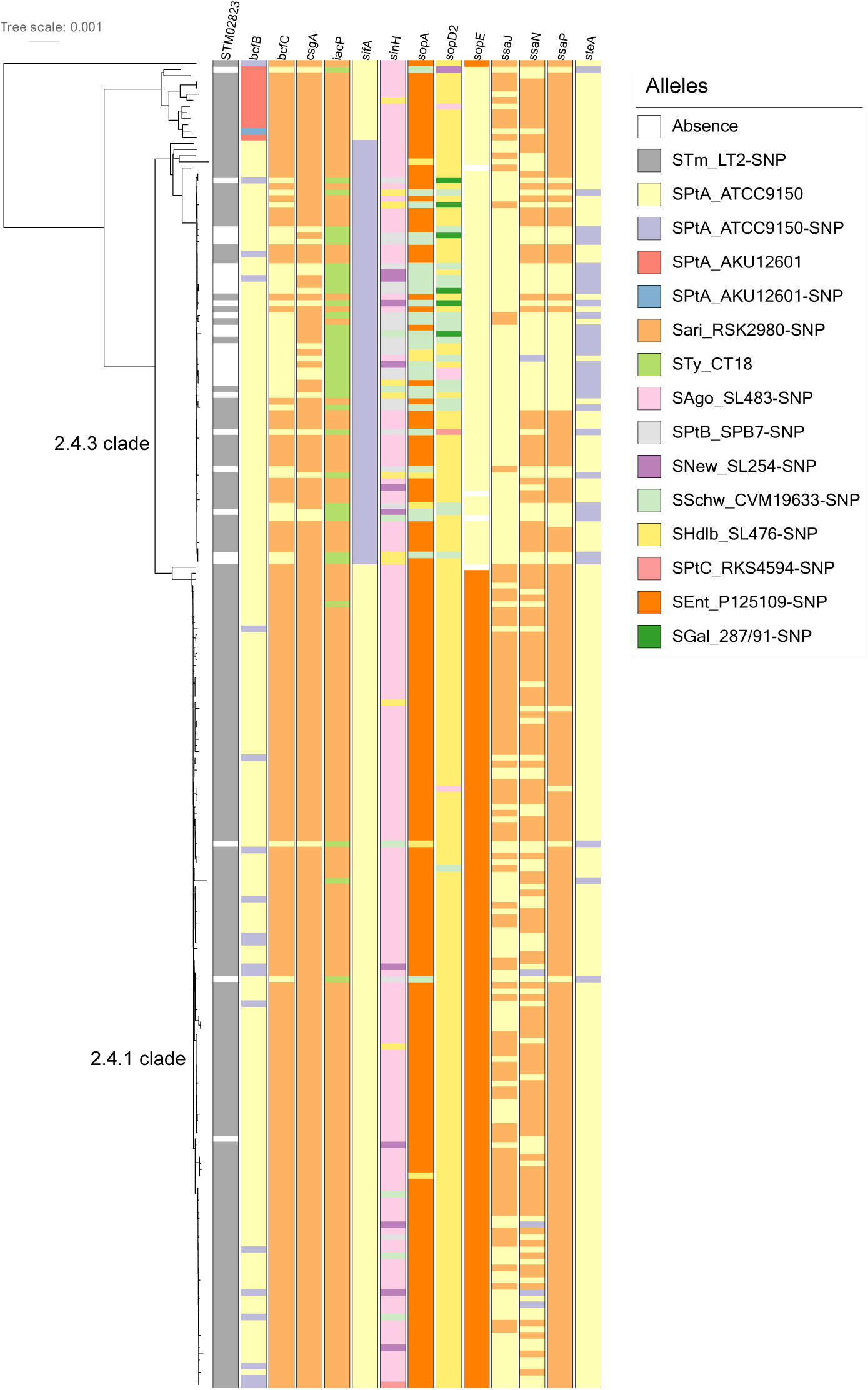
Genetic diversity in virulence factors in the Nepali *S.* Paratyphi A population. Virulence factors with genetic diversity in >20% of isolates as identified using the Virulence Factor Database. Alleles identified are shown in the key (Serotype_strain ID), SNP indicates a mismatch in the allele. STm, *S.* Typhimurium; SPtA, *S.* Paratyphi A; Sari, *S.* Arizona; STy, *S.* Typhi; SAgo, *S.* Agona; SPtB, *S.* Paratyphi B; SNew, *S.* Newport; SSchw, *S.* Schwarzengrund; SHdlb, *S.* Heidelberg; SPtC, *S.* Paratyphi C; SEnt, *S.* Enteritidis; SGal, *S*. Gallinarum.

## Discussion

The control of enteric fever caused by *S.* Paratyphi A is impacted by AMR, poor hygiene, and sanitation measures, as well as the lack of a prophylactic vaccine and a robust framework for sustained surveillance that would allow investigation of population dynamics. Here, we assessed disease dynamics and defined the *S.* Paratyphi A population structure at the genomic level in the highly endemic Kathmandu valley, Nepal, by performing WGS, utilising a recently developed genotyping scheme and associated metadata.

The population of *S.* Paratyphi A studied here comprised of isolates spanning a 9-year period (2005– 2014). The median age of *S.* Paratyphi A patients was 20 years, in line with identified at-risk populations [51]. Carriers were older in age and mostly females, in accordance with associated risk factors for gallbladder removal [3]. Overall, the rate of *S.* Paratyphi A isolation peaked in the summer months, consistent with previous observations regarding the seasonality of enteric fever, and may be associated with increased rainfall and flooding, and/or warmer temperatures, which may promote water contamination [23,52]. Contamination of water sources with coliforms of typhoidal *Salmonella* has been reported in Nepal [47], which may explain the higher counts of *S*. Paratyphi A-infected patients using municipal water irrespective of the treatment method of choice. In addition, the majority of *S.* Paratyphi A in Kathmandu was punctuated by mutations in *gyrA* associated with reduced susceptibility to fluoroquinolones, which was supported by the MIC values of ciprofloxacin. These data further highlight the need for improved access to clean water in endemic areas.

We have very limited understanding of the role of carriers in shaping the population of *S*. Paratyphi A. Although the number of isolated organisms was small, we found a considerable genetic diversity within the carrier isolates, as suggested by the number of genotypes assigned to this group of organisms. The genetic variation observed between *S.* Paratyphi A from enteric fever and those from carriers was associated with the assigned genotype. We found one carrier isolate belonging to genotype 1.1 which was very distant from the rest of the population, differing by a markedly high number of SNPs from all other organisms in this study. This genetic distance from other circulating genotypes serves also as an indication of the length of carriage time in this patient. Importantly, we found carrier isolates to not be restricted to a single genotype and several carrier isolates belonged to genotypes that included several acute isolates circulating in the area. Specifically, carriers of genotype 2.4.1 had many common characteristics with acute isolates, such as antimicrobial susceptibility profiles and mutations in *gyrA* and *rmlB*, suggesting not only that 2.4.1 has been circulating within the Nepali population for some time, but also that carriers reflect the general circulating population. Gallbladder carriage in *S.* Typhi has been suggested to be responsible for enriching the genetic diversity in the circulating population in such a way that carrier isolates show increased numbers of SNPs compared to acute even within the same genotype [4]. In contrast, we did not make such an observation here, suggesting that either these carriers have been infected comparatively recently and the bacterial organisms have not undergone adaptive mutations, or that sustained person-to-person transmission may be taking place. Importantly, chronic carriers have limited contribution to the genetic diversity in the population and genotypes that were identified only in the carrier isolates in this collection did not reappear.

Further exploring the transmission dynamics of the *S.* Paratyphi A population, we observed spatiotemporal clusters incorporating multiple genotypes, indicative of acute transmission of *S*. Paratyphi A in Kathmandu. Previous case-control studies in Indonesia and Nepal also found that acute transmission plays an important role in *S*. Paratyphi A infection with flooding and contaminated street food being important risk factors, whereas contaminated drinking water was more associated with *S*. Typhi infections [9,53]. Noticeably, all these localized *S*. Paratyphi A outbreaks occurred during monsoon months between May and September corresponding to the seasonal distribution of enteric fever in Asia. As mentioned earlier, there is a high degree of faecal contamination in the water supply in correlation with increased rainfall [47]. Only half of the patients in this study had access to municipal water, further supporting that *S*. Paratyphi A infection spreads from a focal point by direct transmission already suggested in our early studies [46].

Outbreaks of enteric fever caused by *S*. Paratyphi A are not uncommon. We recently reported an outbreak of *S*. Paratyphi A in Vadodara, India, which was likely associated with environmental contamination and a reduction in hygiene standards [34]. Additionally, an outbreak of *S*. Paratyphi A infection caused by isolates belonging to the same subclade originating from Cambodia spread to several provinces and sickened many returning travellers [15]. Although the source remained undetermined, contaminated water or food is the most reasonable cause that was then facilitated by acute person to person transmission. It is, therefore, essential that preventative measures including improvement of food safety, water quality and hygiene are implemented to control *S*. Paratyphi A dissemination. Considering, however, that these measures are unlikely to serve as an imminent solution in Nepal, the development of vaccines against *S*. Paratyphi A or a bivalent vaccine protecting against both serovars Typhi and Paratyphi A is of high priority.

Our global *S.* Paratyphi A population analysis suggested that the genotypes observed in Kathmandu in 2005–2014 were circulating in neighbouring countries in the Indian subcontinent and were possibly introduced to Nepal. Interestingly, we observed a clonal expansion of genotype 2.4.3 that spread across Kathmandu and quickly overtook genotype 2.4.1 during the time frame studied here. Genotype 2.4.3 was closely related to genotypes 2.4.2 and 2.4.4 in the neighbouring countries, which appeared in later years, suggesting an exchange of genotypes that circulate in the subcontinent. While we do not currently have data on the prevalent genotype(s) post 2014, it is likely that there are cycles of clonal expansion that are driven by environmental pressures. It remains unclear whether the genotype expansion observed in Nepal was due to successful local clonal expansion or a result of strain introduction from neighbouring countries. Whether the local phylogenetic dynamics of *S*. Paratyphi A in Nepal, namely dominance of a single genotype over time, has also occurred elsewhere in South Asia warrants further investigation.

The lower average SNP difference within genotype 2.4.3 compared to 2.4.1 organisms suggests recent transmission and rapid clonal expansion, that led to the replacement of other genotypes within a four-year period. Our data indicate that, while reduced susceptibility to fluoroquinolones is important for the persistence of successful genotypes of *S*. Paratyphi A, DNA gyrase mutations are present in most genotypes, both Nepali and global, and occurred independently due to increased use of fluoroquinolones, and therefore the clonal expansion of 2.4.3 in Nepal was not solely due to AMR. *S*. Paratyphi A is highly endemic in Nepal, suggesting that entering an immunologically naïve population is possibly not the main driver for the 2.4.3 clade expansion, unless this replacement was associated with subtle changes in antigenicity in these organisms. For example, mutations in the O-antigen *rmlB* and *wzx* genes were associated with specific genotypes (2.4.1 and 2.4.3, respectively). RmlB is essential for rhamnose biosynthesis, which is one of the O-antigen sugars, and Wzx translocates the synthesised O-units across the inner membrane [54]. It is possible that subtle changes on the O-antigen may interfere with antibody recognition and thus immune protection. Given that aspects of O-antigen structure have been shown to directly affect interactions with factors of the immune system [55,56], it is imperative to investigate whether these genetic changes can have a phenotypic impact that may affect interactions with the host. We additionally detected increased genetic variation between 2.4.3 and 2.4.1 *S*. Paratyphi A organisms in several SPI-1 and SPI-2 genes. These pathogenicity islands encode for two type 3 secretion systems (T3SS), which are essential for virulence by mediating entry (SPI-1) and survival (SPI-2) within host cells [57]. Genetic differences in the genes encoding several SPI-1 effector proteins (SifA, SopA, SopD2, SopE) that dictate interactions with host cells, such as cytoskeletal changes and immune signalling [57], in combination with changes in SPI-2 structural components (SsaJ/N/P) that may affect SPI-2 T3SS stability and thus intracellular survival, may have been central to the success of genotype 2.4.3 and warrant further investigation. It is likely that the combined genetic diversity in virulence factors has contributed to the expansion of genotype 2.4.3.

In summary, the *S*. Paratyphi A population in Nepal is largely clonal marked by mutations in DNA gyrase and increased genetic variation in other virulence-associated genetic loci. We observed an unusual clonal expansion and replacement by a single genotype, which may be attributed to both reduced susceptibility to fluoroquinolones and to the genetic variation in virulence genes. Rapid dissemination within the local population and diversification of the dominant clones have resulted in short-term spatiotemporal clusters of various genotypes indicating the importance of direct transmission of *S*. Paratyphi A in this setting. We further suggest that in a highly endemic setting, chronic carriage has a limited contribution to disease transmission and maintenance of the genetic diversity.

## Data Availability

All supporting data, code, methods, and accession numbers have been provided within the article or through supplementary data files. Two supplementary tables with all accession numbers are available.

## Acknowledgements

We wish to thank all patients participating in the contributing clinical studies and the diagnostics services helping collecting samples.

---

**Figure S1.**
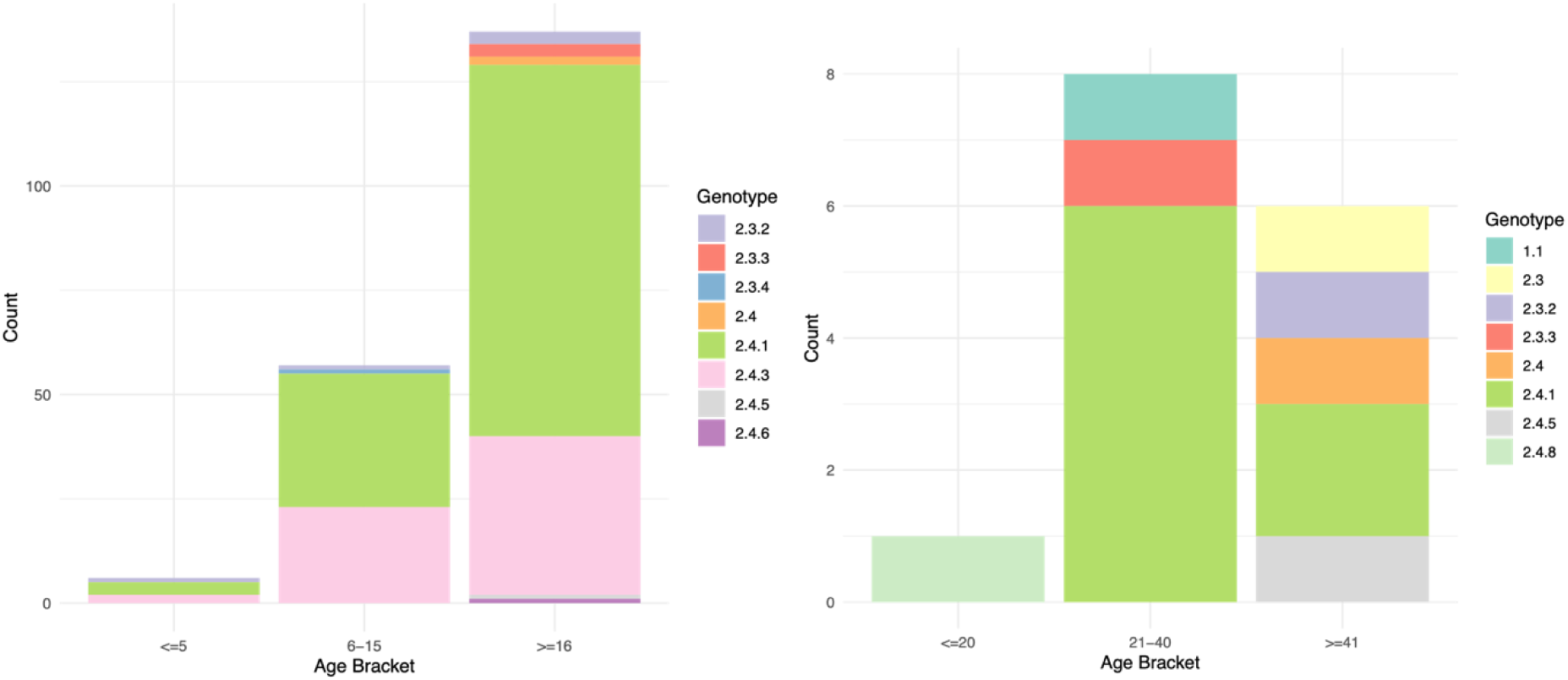
*S.* Paratyphi A genotype distribution by age of acute enteric fever patients (left) and chronic carriers (right). Bar charts show counts of patients in each age bracket.

**Figure S2.**
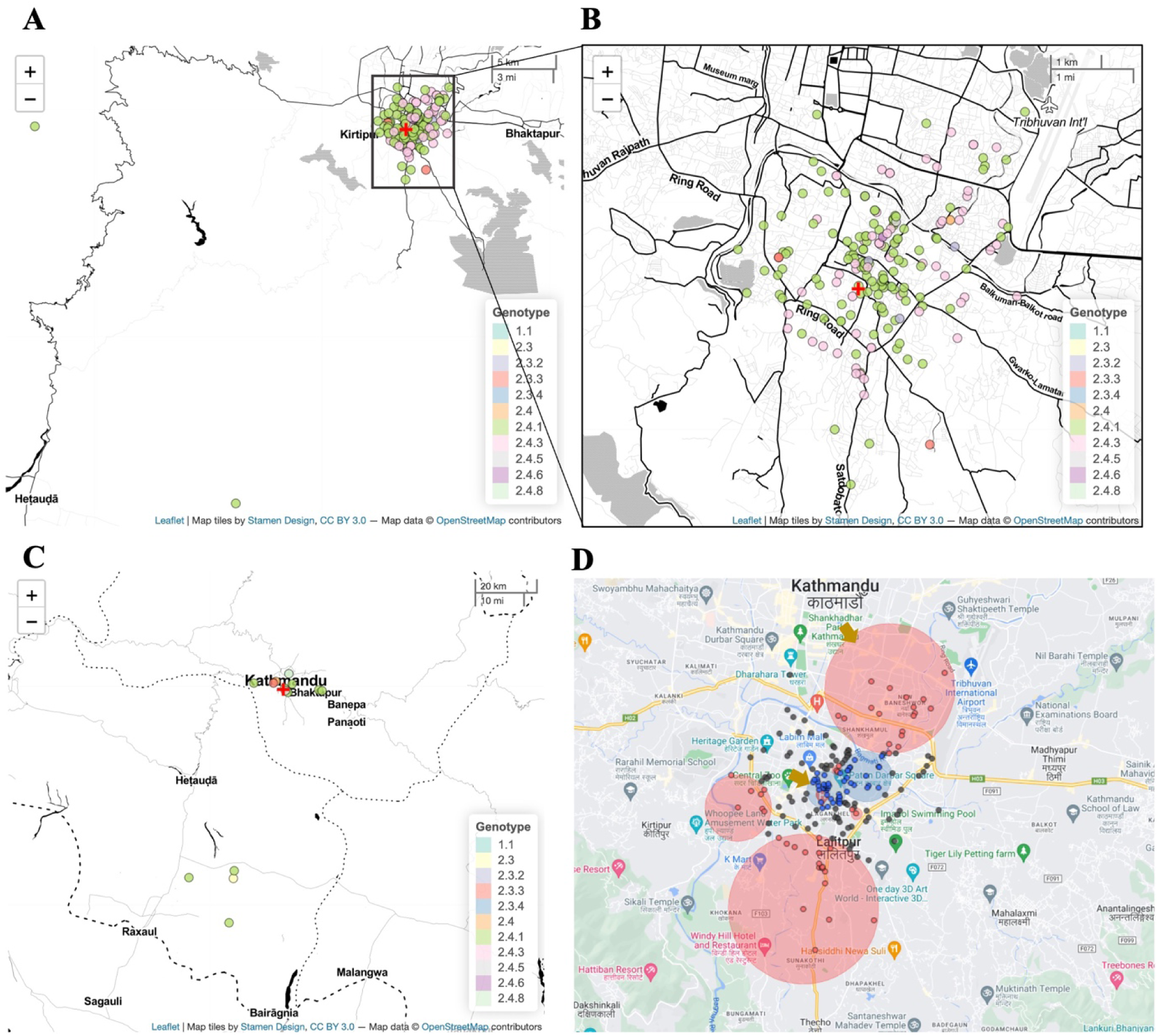
Geographical distribution of *S.* Paratyphi A genotypes. (A) Genotypes of acute *S*. Paratyphi A organisms with (B) a zoomed in inset of the majority of cases. (C) Genotypes of carrier *S.* Paratyphi A organisms. (D) Spatiotemporal clustering of *S.* Paratyphi A with red circles showing high rate clusters and the blue circles indicating low rate clusters (statistically significant clusters, *p*<0.05, are indicated with yellow arrows). Red cross indicates the Patan Hospital.

**Figure S3.**
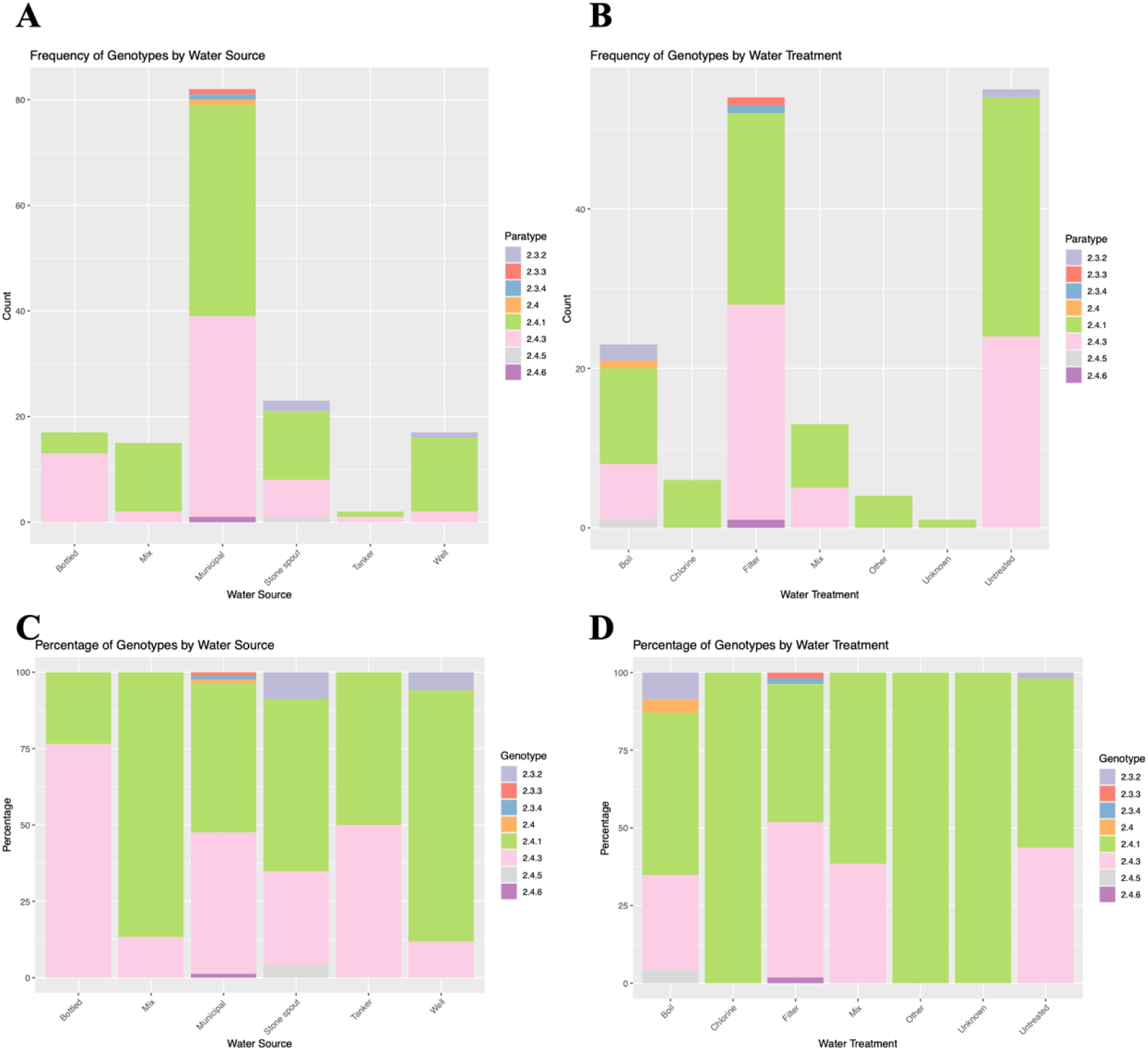
Genotype distribution with water source and water treatment from 156 enteric fever patients. Stacked bar plots showing the (A-B) count and (C-D) percentages of each genotype for each associated water source (A-C) and water treatment (B-D).

